# Confirmation of the *MIR204* n.37C>T heterozygous variant as a cause of chorioretinal dystrophy variably associated with iris coloboma, early-onset cataracts and congenital glaucoma

**DOI:** 10.1101/2023.02.09.23284763

**Authors:** J. Jedlickova, M. Vajter, T. Barta, G. Black, J. Mares, M. Fichtl, B. Kousal, L. Dudakova, P. Liskova

## Abstract

Four members of a three-generation family with early-onset chorioretinal dystrophy were shown to be heterozygous carriers of the n.37C>T in *MIR204*. The identification of this previously reported pathogenic variant confirms the existence of a distinct clinical entity caused by a sequence change in *MIR204*. The chorioretinal dystrophy was variably associated with iris coloboma, congenital glaucoma, and premature cataracts extending the phenotypic range of the condition. In silico analysis of the n.37C>T variant revealed 713 novel targets. Additionally, family members were shown to be affected by albinism resulting from biallelic pathogenic *OCA2* variants.

## INTRODUCTION

Inherited retinal degenerations and major structural developmental eye disorders (microphthalmia, anophthalmia, and coloboma) are a common cause of visual impairment in childhood. One of the genes reported to be implicated in the development of these disorders is *MIR204* (MIM *610942). Previously the n.37C>T variant, in the seed region of MIR204, was found in 6 members from a five-generation family of white British descent and the associated disease was termed “retinal dystrophy and iris coloboma with or without congenital cataract” (RDICC, MIM **#**616722).^1^

MicroRNAs (miRNAs) are short (20-24 nt) noncoding RNAs involved in post-transcriptional regulation most commonly leading to translational inhibition or degradation of the target mRNA.^2^ MIR204 is necessary for normal eye development including photoreceptor differentiation and function. Morpholino-mediated knockdown of miR-204 in medaka fish resulted in microphthalmia, abnormal lens formation, and altered dorsoventral patterning of the retina with optic fissure coloboma.^1,3^

In this study, we report a three-generation family with the occurrence of early-onset chorioretinal dystrophy variably associated with iris coloboma, premature cataracts, and congenital glaucoma in four members. Heterozygous variant n.37C>T in *MIR204* was detected. In addition, molecular investigation resolved the presence of albinism in the spouse of the proband and his three children by identifying *OCA2* pathogenic variants, including one daughter with dual diagnosis.

## METHODS

### Sample collection and clinical examination

The study followed the tenets of the Declaration of Helsinki and was approved by the Ethical review board of General University Hospital in Prague (reference number 412/20 S-IV). All investigated subjects or their legal representatives provided informed consent with examination and publishing of the results prior to the inclusion into the study.

Ocular examination included best corrected visual acuity (BCVA) measurements using Snellen charts with extrapolation to decimal values, slit-lamp evaluation including fundoscopy and static automated perimetry (M-700, Medmont International, Vermont, Australia). Colour fundus photography was obtained using Clarus 700 and/or FF 450 plus IR (Carl Zeiss Meditec AG, Jena, Germany). Spectral domain optical coherence tomography (SD-OCT) was performed using Spectralis (Heidelberg Engineering GmbH, Heidelberg, Germany). Severity of foveal hypoplasia (grades 1-4) was assessed using morphological findings obtained by SD-OCT according to a previously described scheme.^4^

### DNA extraction and variant description

DNA was extracted using Gentra Puregene™ Blood Kit (Qiagen, Hilden, Germany) or Oragene saliva kit (Oragene OG-300, DNA Genotek, Canada) according to the manufacturer’s instructions. Variant description followed the Human Genome Variation Society (HGVS) guidelines.^5^ The pathogenicity of the detected variants was evaluated according to the American College of Medical Genetics and Genomics (ACMG) and the Association for Molecular Pathology recommendations.^6^

### Sequencing

Exome sequencing was performed in the proband’s daughter (individual III:4, Fig. 1) and spouse (individual II:3, Fig. 1) using SureSelect Human All Exon V6 Kit (Agilent, Santa Clara, California, USA), sequenced on a NovaSeq 6000 instrument (Illumina, San Diego, CA) with 150 bp paired-end reads. Sequence reads were aligned against the hg19 version of the human genome, using the Burrows-Wheeler Aligner (http://bio-bwa.sourceforge.net/). Variant calling was performed with Genome Analysis Toolkit (version 4.2.6.1).^7^ Only rare variants (i.e. minor allele frequency ≤ 0.001 as per gnomAD v2.1.1, v3.1.2)^8^ were further evaluated for potential pathogenicity.

**Figure 1:**
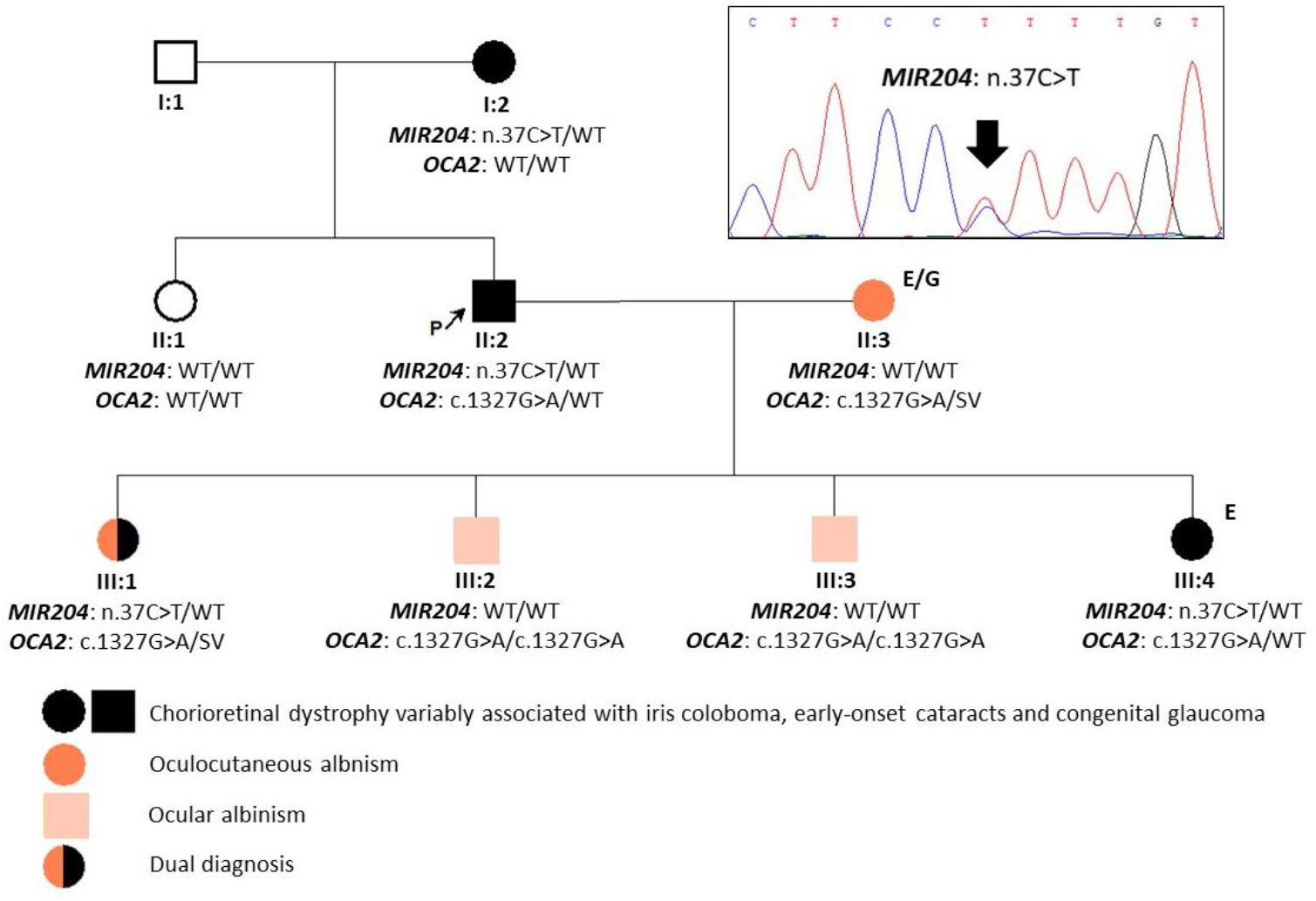
Pedigree of the family and segregation of identified pathogenic variants in *MIR204* and *OCA2*. NR_029621.1 and NM_000275.3 were taken as the reference sequences. E - exome sequencing; G - genome sequencing; SV - structural variant (*OCA2* “143kb;184kb CxSV” allele); WT - wild type.

Genome sequencing was performed in the proband’s spouse with the aim to search for the second pathogenic variant in *OCA2*. NEBNext Ultra DNA library Prep kit with 150 bp paired-end reads (Illumina) was used together with the Illumina NovaSeq 6000 sequencing platform. Manual inspection of genome data in Integrative Genomics Viewer (IGV),^9^ allowed for designing a set of primers to Sanger sequence the breakpoints of a detected structural rearrangement in *OCA2*. Sanger sequencing was also used to track the segregation of pathogenic variants. All primers used are listed in Supplementary Table 1 and Supplementary Figure 1.

### miRNA target analysis

To investigate the putative functional consequences of the n.37C>T variant on its target genes, we applied miRNA target prediction tool miRDB (version 6.0).^10,11^ Both sequences WT hsa-miR-204-5p (uucccuuugucauccuaugccu) as well as n.37C>T hsa-miR-204-5p (uuccuuuugucauccuaugccu) were submitted to the miRDB. The generated list of genes including target scores for each variant (i.e. was used to create a heatmap (R version 4.1.2 and pheatmap package). All the predicted targets have prediction scores between 50 and 100, which are assigned by the computational algorithm. A predicted target with score >80 is considered likely to be real. If the score is <60, it is recommended to provide additional supporting evidence.

The functional annotation was generated by the Database for Annotation, Visualization and Integrated Discovery (DAVID tool, version 2021)^12^ using gene ontology annotation. Only significant targets (p<0.001) were selected for the creation of the heatmap. The hairpin structure was analysed using the RNAfold tool (http://rna.tbi.univie.ac.at/cgi-bin/RNAWebSuite/RNAfold.cgi).

## RESULTS

### MIR204 n.37C>T causes a distinct chorioretinal dystrophy

We have identified a family with four members in three generations affected by chorioretinal dystrophy variably associated with iris coloboma, early-onset cataracts, and congenital glaucoma.

Exome sequencing showed that the youngest daughter of the proband (individual III:4, Fig. 1) is a heterozygous carrier of the n.37C>T in *MIR204* (NR_029621.1), which has been previously reported in one family.^1^ Segregation analysis confirmed the presence of the n.37C>T in *MIR204* in three other affected family members, i.e. the proband, his mother, and his oldest daughter (Fig. 1). The variant was classified as pathogenic according to the ACMG criteria: PS3_Strong (functional studies show a damaging effect), PS4_Supporting (prevalence of the variant in affected individuals is significantly increased compared with controls, 2^nd^ independent occurrence), PM1_Supporting (variant is located in a well-established functional domain), PM2_Supporting (extremely low-frequency in population databases), PP1_Moderate (co-segregation with a disease in multiple affected family members, 5-6 meiosis), PP4_Moderate (patient’s phenotype or family history is highly specific for a disease with a single genetic aetiology).

No rare variants evaluated as possibly pathogenic were identified in other genes associated with developmental glaucoma and ocular coloboma in exome data of individual III:4 [PanelApp Ocular coloboma (Version 1.46) and Glaucoma (developmental) (Version 1.42)].^13^

### Additional molecular genetic findings

The spouse of the proband was found to be a compound heterozygote for the pathogenic missense variant c.1327G>A p.(Val443Ile) in *OCA2* (NM_000275.3) and a large complex pathogenic rearrangement, previously characterised in detail by Loftus et al.^14^ In brief, a 143 kb inverted segment is reintroduced in intron 1 followed by an additional 184 kb deletion across the same region, restoring exons 3–19 of *OCA2* to a copy-number neutral state. This variant is defined by the HGVS nomenclature as NC_000015.9:g.[28337021_28339403delins[CCTGGTTGTAGGTCTAACCTGGTTAGAATCA;28143225_28285967inv; C];[28119923_28303785del]. Loftus et al. suggested denoting this structural variant (SV) as “143kb;184kb CxSV” allele. It has been predicted to generate an mRNA transcript with novel splicing between exons 2 and 20, thus resulting in premature truncation and a functionally null *OCA2* allele p.(Ser77Hisfs*7).^14^ The SV has been repeatedly observed in compound heterozygosity with pathogenic variants in patients diagnosed with albinism.^14^

Segregation analysis (Fig. 1) revealed that the proband is also a heterozygous carrier of the c.1327G>A variant in *OCA2* which explained pseudodominance for albinism in this particular family. The oldest daughter was shown to be a compound heterozygote for the c.1327G>A inherited from the proband and the pathogenic *OCA2* SV inherited from her mother, in addition to harbouring the n.37C>T in *MIR204*. Both sons had the *OCA2* c.1327G>A in a homozygous state.

### Clinical features in the extended family

The mother of the proband (individual I:2, Fig. 1) with no family history of ocular disease had bilateral retinal lesions, which were according to the available past medical records considered as a sequela of retinal toxoplasmosis in first decade of life. Due to gradual visual functions worsening, she started to attend school for the visually impaired in the adolescence. In her 30s she was diagnosed with a left eye cataract which was removed two years later. Upon clinical examination performed in her 70s, the patient was completely blind with no light perception. There was also cataract in the right eye, iris coloboma in the left eye, and advanced bilateral chorioretinal dystrophy (Table 1, Fig. 2A-D).

**Table 1:** Clinical findings of affected individuals in the examined family.

| ID | Primary diagnosis | Age | BCVA RE (refraction) | BCVA LE (refraction) | Anterior segment | Posterior segment | Other | Surgical management |
| --- | --- | --- | --- | --- | --- | --- | --- | --- |
| I:2 | CRCCG | 30s | LP | 0.1 (-0.5 DS) | LE iris coloboma, BE early onset cataract | BE chorioretinal atrophy with central and peripheral RPE clumping | Night blindness, BE nystagmus | LE cataract surgery in her 30s |
|  |  | 70s | NLP | NLP |  |  |  |  |
| II:2 | CRCCG | 10s | 0.04 (-3.0 DS) | 0.04 (-3.5 DS) | BE persistent pupillary membrane, BE iris coloboma, BE early onset cataract | BE chorioretinal atrophy with central and peripheral RPE clumping | Night blindness, BE nystagmus | RE cataract surgery in the 20s |
|  |  | 15s | 0.04 (plan) | 0.03 (plan) |  |  |  |  |
|  |  | 25s | CF | LP |  |  |  |  |
|  |  | 40s | LP | LP |  |  |  |  |
| II:3 | OCA2 (oculocutaneous) | 40s | 0.32 (+1.5 DS/-1.0 DC) | 0.5 (+2.5 DS/-2.0 DC) | BE iris translucency | BE fundus hypopigmentation, BE foveal hypoplasia (grade 4) | BE nystagmus, white hair and very light-colored skin | None |
| III:1 | CRCCG+OCA2 (oculocutaneous) | 15s | 0.04 (plan) | 0.063 (plan) | BE iris translucency | BE fundus hypopigmentation, chorioretinal atrophy and irregular RPE distribution but no clumping, BE foveal hypoplasia (grade 4) | BE nystagmus, BE visual field constriction, white hair and very light-coloured skin | None |
| III:2 | OCA2 (ocular) | 10s | 1.0 (plan) | 1.0 (plan) | Normal | BE foveal hypoplasia (grade 1) | None | None |
| III:3 | OCA2 (ocular) | 10s | 1.0 (plan) | 1.0 (plan) | Normal | BE foveal hypoplasia (grade 1) | None | None |
| III:4 | CRCCG | 5-10 y | 0.06 (-3.0 DS) | LP (-3.0 DS) | BE iris coloboma, LE band keratopathy | BE fundus hypopigmentation, BE atrophic lesion in the macula demarcated by pigmentary deposits | BE congenital glaucoma, BE nystagmus | BE CPC <1 y, RE TE <4 y, LE CPC 2x between 3-5 y, LE EDTA chelation <5 y |

**Abbreviations:** BCVA - best corrected visual acuity; BE - both eyes; CF - counting fingers; CPC - cyclophotocoagulation; CRCCG - chorioretinal dystrophy variably associated with iris coloboma, early-onset cataract and congenital glaucoma; D - diopter; DC - diopter cylinder; DS - diopter sphere; EDTA - ethylenediaminetetraacetic acid; FH - foveal hypoplasia; LE - left eye; m - months; NLP - no light projection; RE - right eye; RPE - retinal pigment epithelium; TE - trabeculectomy; y - year

**Figure 2:**
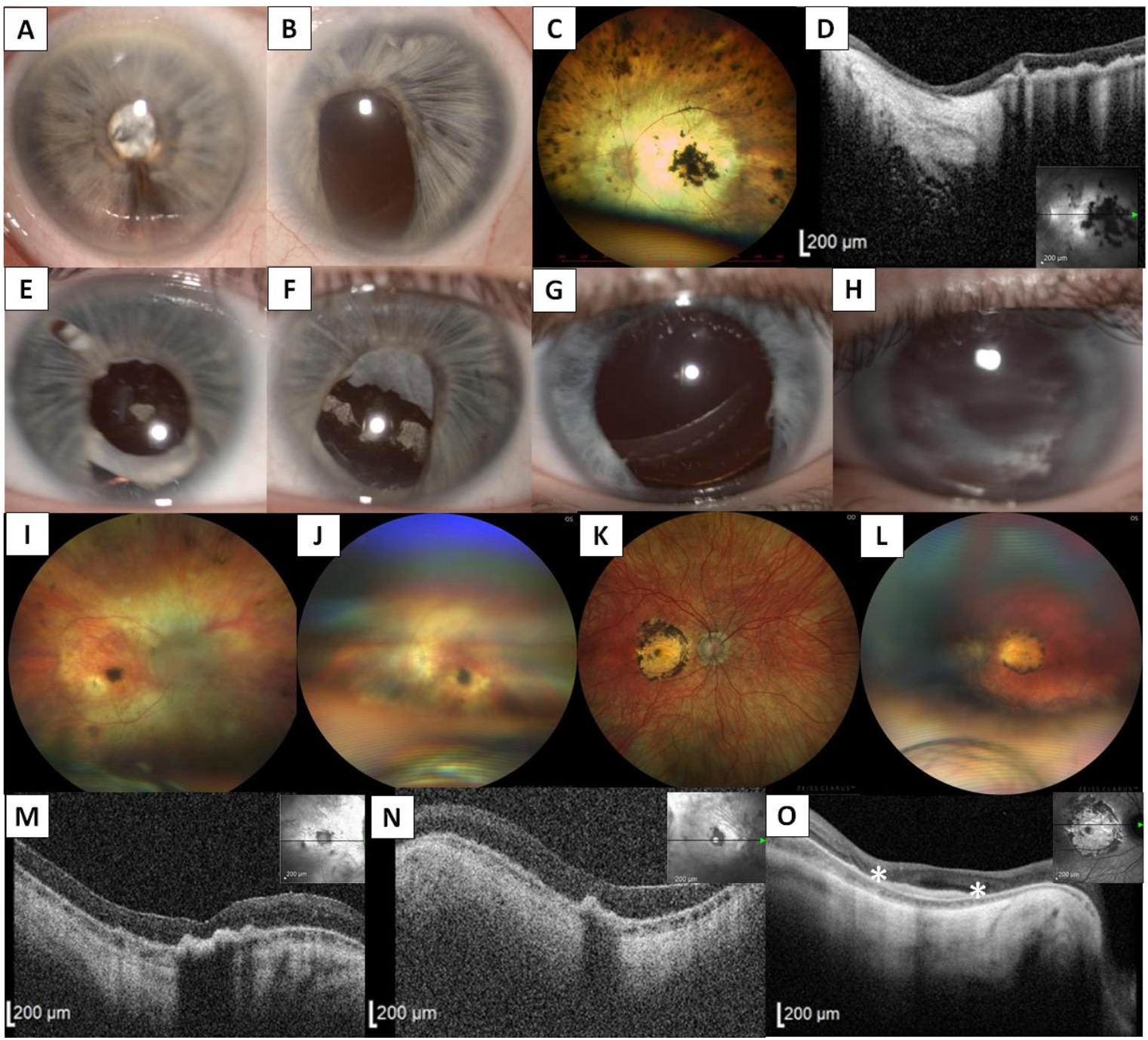
Clinical findings in three family members with dominantly inherited chorioretinal dystrophy variably associated with iris coloboma, congenital cataract, and glaucoma. Anterior segment photography of individual I:2 showing iris hypoplasia and intumescent cataract in the right eye (A), and iris hypoplasia with coloboma and aphakia in the left eye (B). Left eye fundus photograph showing advanced chorioretinal atrophy with pigment clumping in both the macula and retinal periphery (C) and corresponding transfoveal horizontal SD-OCT scan documenting severe atrophy and thinning of the neurosensory retina and hyperplasia of the retinal pigment epithelium (D). Anterior segment images of the proband (II:2) showing iris hypoplasia and coloboma in the right (E) and left eye (F), and in individual III:4 documenting iris hypoplasia and coloboma in the right (G) and left eye (H). Pale optic discs and advanced chorioretinal dystrophy manifesting as pale fundus with attenuated vessels, atrophic lesion with pigmentary mottling in the macula, and occasional pigmentary deposits in the periphery in the right (I) and left eye (J) in the proband. In comparison, his daughter (individual III:4) had a round lesion of chorioretinal atrophy with pigmented edges in the macula disrupting the entire fovea in the right (K) and left (L) eye. Corresponding transfoveal horizontal SD-OCT scans of the right (M) and left (N) eye of the proband document chorioretinal atrophy, involving all retinal layers and subfoveal hyperplasia of the retinal pigment epithelium. SD-OCT in the daughter also shows chorioretinal atrophy and subfoveal hyperreflective lesions corresponding to the areas of subretinal fibrosis in the right eye (present between the asterisks) (O). The inserts in D, M, N, O provide a cross-sectional plane. Retinal imaging quality is decreased in both the proband and his daughter due to nystagmus and anterior segment findings.

Clinical notes of the proband (individual II:2, Fig. 1) documented complaints about nyctalopia and bilateral retinal dystrophy misdiagnosed as congenital toxoplasmosis in adolescence. The manifestation of cataract is unknown, however available medical documentation described it as congenital. Bilateral cataract surgery was performed in his early 20s. Serial measurements of BCVA showed gradual decline, with counting fingers in the right eye and light projection in the left eye in the third decade of life (Table 1). When examined by the authors, in his early 40s, there was a bilateral advanced chorioretinal atrophy (Fig. 2, Table 1).

The proband had four children. The youngest one, in early school age at the time of clinical evaluation (individual III:4, Fig. 1), was noted to suffer from congenital glaucoma for which she had been treated elsewhere. She underwent several antiglaucoma procedures in both eyes (Table 1). Our examination revealed bilateral macular atrophic lesions with pigmented edges. In addition, her fundus appeared pale bilaterally. There were no signs of premature cataracts. She also developed band keratopathy in both eyes (Fig. 2G, H). BCVA was 0.06 and light projection in the right and left eye, respectively.

The oldest child, (individual III:1, Fig. 1), was referred in adolescence with oculocutaneous albinism. The clinical evaluation showed markedly low BCVA; 0.04 in the right eye and 0.063 in the left eye, and advanced visual field constriction. SD-OCT revealed absent foveal pit/depression and chorioretinal atrophy (Fig. 3). Fundus was hypopigmented bilaterally but there were no focal lesions or pigment deposits.

**Figure 3:**
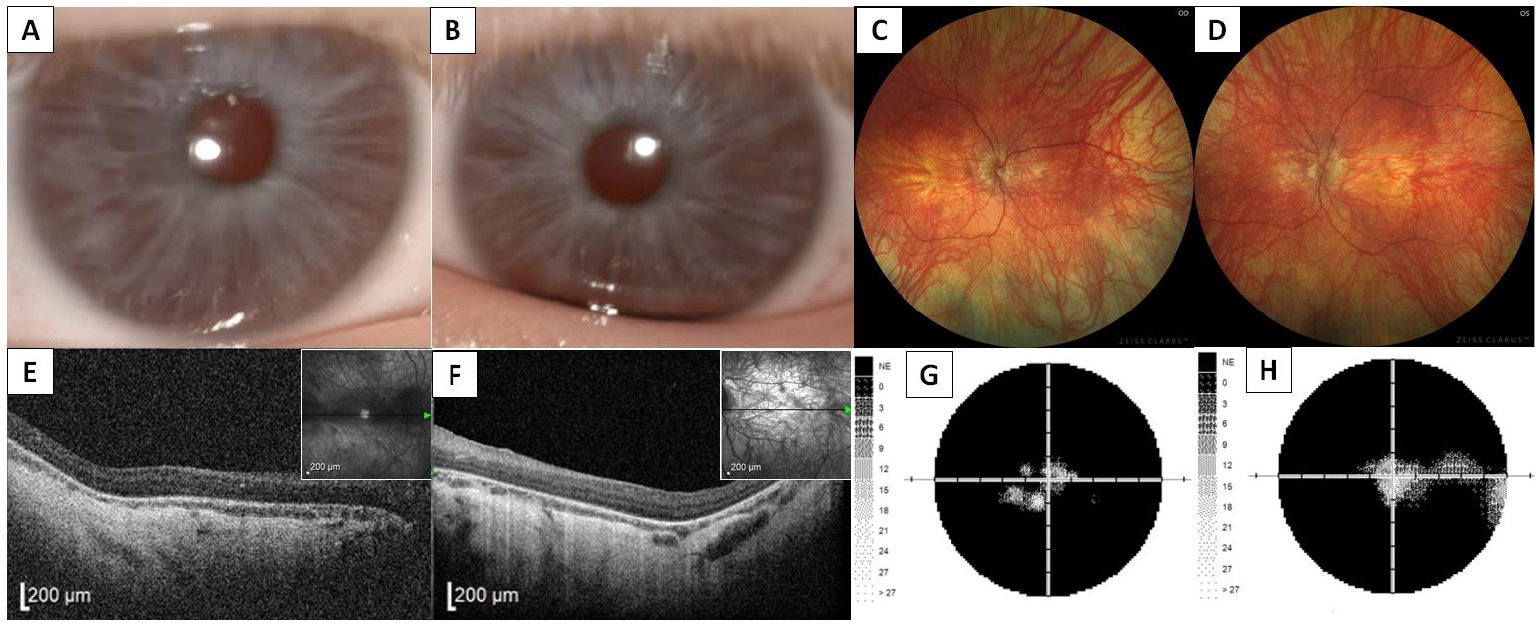
Clinical findings in individual III:1 with dual diagnosis of dominantly inherited chorioretinal dystrophy variably associated with iris coloboma, congenital cataract and glaucoma and oculocutaneous albinism. Anterior segment photography documenting iris translucency, no iris coloboma and no cataract in the right (A) and the left eye (B), respectively. Right (C) and left eye (D) wide-field fundus photography showing marked diffuse hypopigmentation, without pigment clumping. Transfoveal horizontal SD-OCT scans of the right (E) and left (F) eye documenting absence of foveal pit, loss of outer retinal layers and choroidal thinning. Static perimetry revealed concentric visual field constriction with some preserved islands of perception in the right (G) and left eye (H).

The spouse of the proband (individual II:3, Fig. 1) had, similarly to her daughter, typical oculocutaneous albinism characterised by pale skin, hair, eye lashes, iris translucency, fundus hypopigmentation, with corresponding foveal hypoplasia and moderate decrease of BCVA (Supplementary Fig. 2). Two sons of the proband, in the pre-teen and teenage years (individuals III:2 and III:3, Fig. 1), reported no visual symptoms and correspondingly their uncorrected visual acuity was 1.0 in both eyes. Slit-lamp examination did not reveal any anterior segment pathology. There was a discreet fundus hypopigmentation and SD-OCT scans showed bilateral foveal hypoplasia (grade 1) in both children (Supplementary Fig. 3).

Clinical findings of all the affected family members including BCVA at various time points are summarised in Table 1.

### In silico target prediction and functional annotation approach for the n.37C>T in MIR204

Finally, we analysed the impact of the *MIR204* n.37C>T variant on the hairpin structure and function using *in silico* target prediction and functional annotation approach. First, we found no profound changes to the hairpin structure and composition introduced by the n.37C>T (Fig. 4A), presumably having no or little impact on the preferential processing of the -3p or - 5p miRNA strands in the miRNA molecular machinery. The n.37C>T variant gives a rise to the miRNA molecule (n.37C>T hsa-miR-204-5p) that possesses a change in its seed sequence on the 4th position when compared to wild type (WT hsa-miR-204-5p) (Fig. 4A).

**Figure 4:**
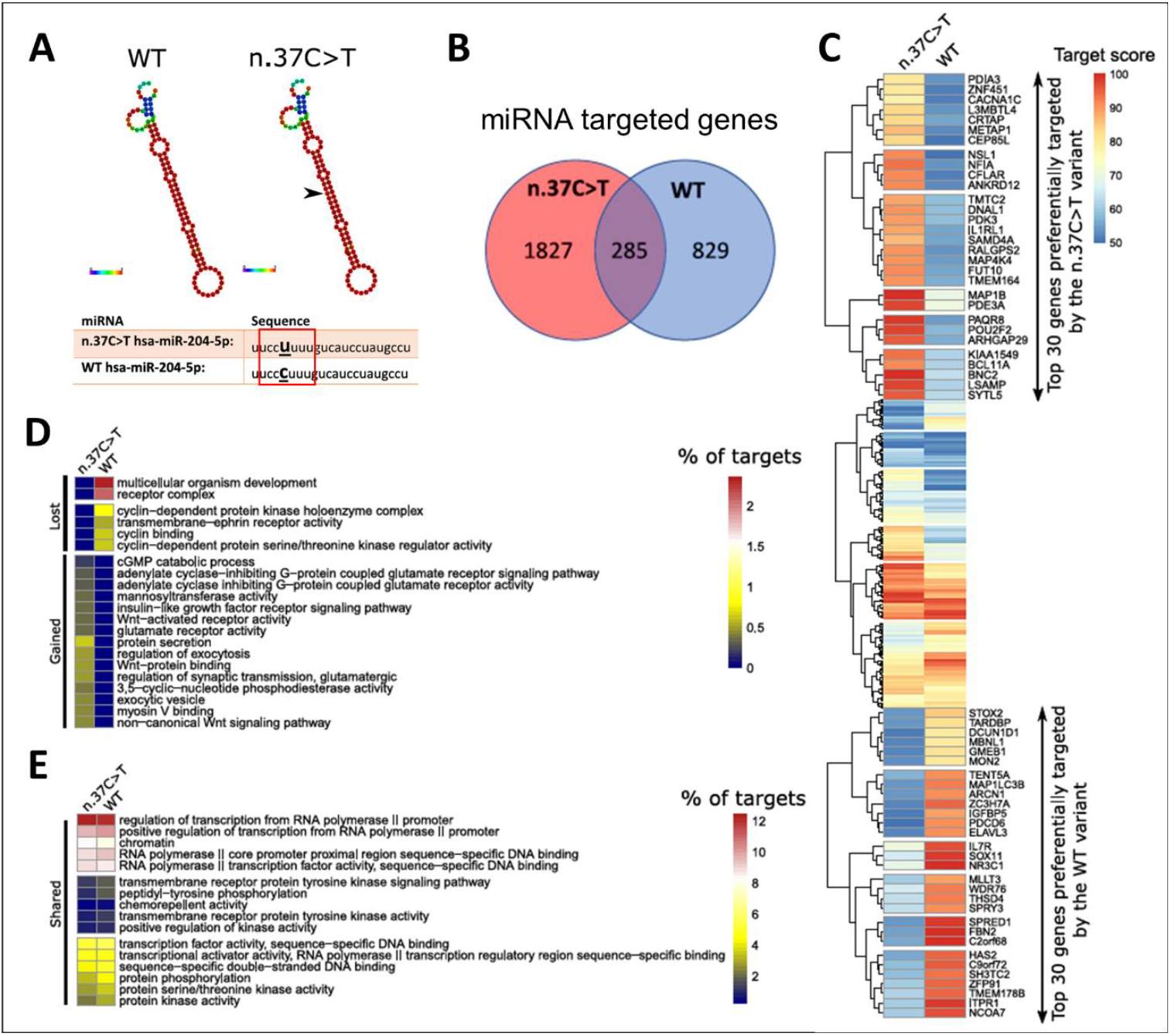
In silico analysis of the *MIR204* n.37C>T variant reveals profound changes to its target mRNAs and suggests a gain-of-function mechanism of *MIR204* variant. Hairpin structure of WT and n.37C>T variant of the *MIR204*, as determined using the RNAfold tool. Black arrow shows the site of the single nucleotide change. Red square marks the seed sequence of *MIR204* (A). Number of predicted targets for both WT as well as n.37C>T variant, as determined using miRDB tool (B). Heatmap shows a comparison of the putative functional consequences of the n.37C>T and WT variant on target genes (C). Functional annotation of target genes of both *MIR204* variants, as determined using DAVID tool (D, E).

Since a change in a seed sequence may have a deep impact on miRNA target recognition, potentially leading to a new gain-of-function mechanism of the mutated miRNA molecule, we also aimed to analyse the effects of the n.37C>T on the predicted hsa-miR-204-5p target genes. The miRNA molecule carrying the pathogenic variant may target 2112 mRNAs, compared to 1114 mRNAs of the WT, with 285 genes shared (Fig. 4B), indicating that the n.37C>T variant not only introduces profound changes to preferential binding to mRNA targets but also binds to novel mRNAs.

To reveal how much the point change affects binding to its targets, we analysed binding scores for each shared mRNA target. The analysis revealed that the n.37C>T pathogenic variant changes the miRNA binding preferences for the majority of shared target genes (Fig. 4C). Given the striking alterations in the target preferences of the n.37C>T variant, we assessed *in silico* its functional consequence on diverse molecular mechanisms. The functional annotation analysis suggests that the n.37C>T variant gains novel abilities to regulate crucial signalling pathways including canonical and non-canonical Wnt pathway, insulin-like growth factor pathway and G-protein coupled glutamate receptor pathway, while it loses the ability to directly regulate cell cycle via cell cycle-dependent protein kinases and cyclins (Fig. 4D). Additionally, the n.37C>T variant shares multiple functional annotation clusters with the WT variant, however, the number of target genes significantly differs (Fig. 4E).

## DISCUSSION

Reported pathogenic variants within miRNAs associated with Mendelian disorders are rare. To date, four have been identified as a cause of human disease.^15,16^ Conte et al. implicated a point variant in miRNA in inherited retinal dystrophy.^1^ In this study, we confirm that the n.37C>T variant in *MIR204* is disease-causing in the heterozygous state.

Clinical examination and available past medical notes of four affected members in the second family identified worldwide showed that the only constant sign of MIR204-associated disease is early-onset chorioretinal dystrophy which we were able to document in a school age child. Interestingly, we observed central macular atrophy surrounded with retinal pigment epithelium (RPE) resembling toxoplasmosis. Clinical notes indicated that similar lesions were also found in the proband and his mother. An atrophic macular lesion with RPE clumping was not detected in the daughter with the dual diagnosis of *MIR204*-associated chorioretinal dystrophy and oculocutaneous albinism; it remains unclear how the intersection of these two genetic defects influences retinal morphology. Nevertheless, in line with the presence of the *MIR204* n.37C>T, her visual function was significantly worse than her mother in her 40s who only had oculocutaneous albinism.

Coloboma was not a constant sign, although it was present in at least one eye of three out of the four individuals carrying the n.37C>T in *MIR204*. The diagnosis of bilateral congenital glaucoma, which was not described in the previously published family with *MIR204* n.37C>T, led us to perform exome sequencing in the affected child. By excluding pathogenic variants in genes associated with congenital glaucoma we propose congenital glaucoma as a rare phenotypical sign of *MIR204*-associated disease, rather than a separate entity.

Changes in seed sequence, which is essential for the miRNA binding to the target mRNA, may lead to binding disruption and/or alterations to premiRNA processing.^16,17^ Notably, the expression of both premiRNA, as well as processed -3p/-5p forms of the *MIR204* n.37C>T variant was not affected when compared to the wild type, suggesting that the mutation does not alter premiRNA processing.^1^ On the other hand, alterations in the seed sequence may result in profound changes to the expression of the miRNA target genes and additionally lead to binding to novel mRNAs representing the gain-of-function mechanism. The n.37C>T variant likely leads to the gain-of-function mechanism by creating novel aberrant recognition sites in genes not normally targeted by miR-204. This scenario is largely supported by the miRNA target and functional annotation analyses performed in this study. Additionally, injection of the n.37C>T *MIR204* variant into medaka fish embryo leads to severe ocular malformations including retinal dystrophy.^1^ Moreover, *MIR204* loss-of-function has been previously linked to microphthalmia, abnormal lens development, eye coloboma, and defects in axon projections of retinal ganglion cells in fish, highlighting the importance of the *MIR204* in eye development and function.^3,18,19^ However, the precise molecular mechanism of the *MIR204*-associated disease remains still unknown.

Another ocular condition running in the family was OCA2 presenting as oculocutaneous form in the spouse and in one of the offspring who had a dual diagnosis. Interestingly, she did not show atrophic lesions in the macula or RPE clumping. Two sons shown to have OCA2 at molecular level presented with normal visual acuity, discrete fundus hypopigmentation and foveal hypoplasia, which was only revealed by chance due to familial investigation.

The *MIR204* belongs to the group of recently characterised retinal-specific miRNAs^20,21^ regulating important processes in the retina including phototransduction cascade and development of RPE, neural retina, ciliary body, and lens. Given the critical role of the *MIR204* in the ocular development and function of the retina, it is not surprising that pathogenic variants in the *MIR204* gene lead to severe eye defects. As the current screening methodology leaves approximately 40% of patients with inherited retinal disorders undiagnosed, identification of the molecular cause is one of the current challenges. In general, the pathogenic role of miRNAs may have been significantly underestimated and could explain part of the missing heritability.^22^

## Data Availability

All data produced in the present study are available upon reasonable request to the authors.

## ACKNOWLEDGEMENTS

Funding: This publication was supported by MH CZ-DRO-VFN64165, NU20-07-00182, and by national funds from the Ministry of Education, Youth and Sports under the European Joint Program for Rare Diseases (Solve-RET 8F20004 No. 825575). PL and LD were also supported by UNCE/MED/007 and JJ by SVV 26367/2017. TB was supported by the Czech Science Foundation (GA21-08182S).

---

**Supplementary Table 1.**
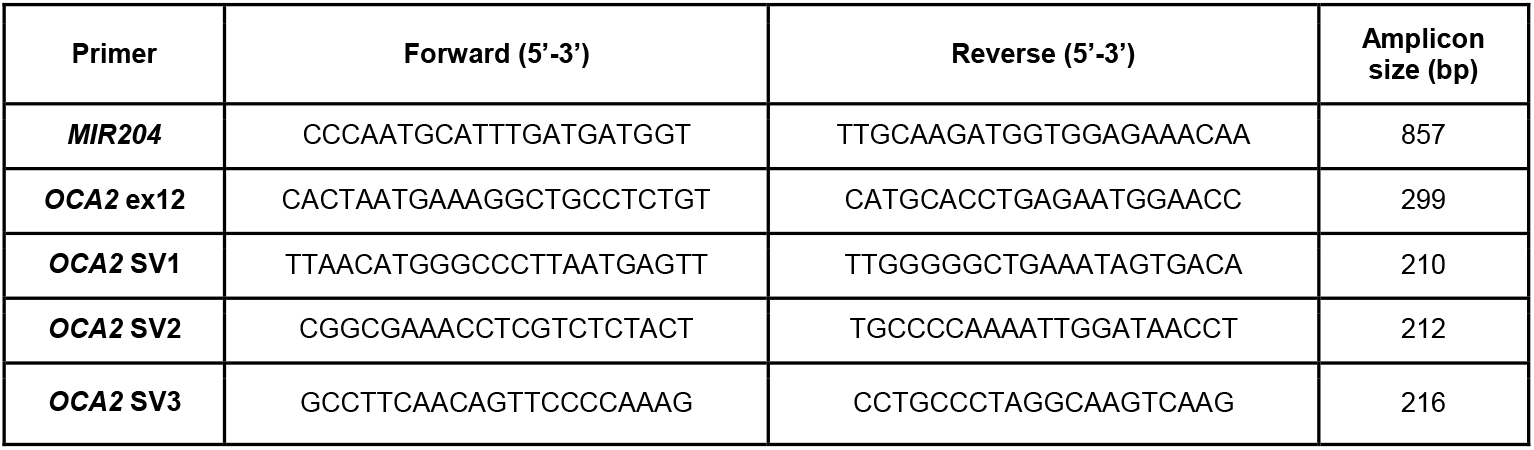
Sequence of primers used for PCR and Sanger sequencing.

**Supplementary Figure 1.**
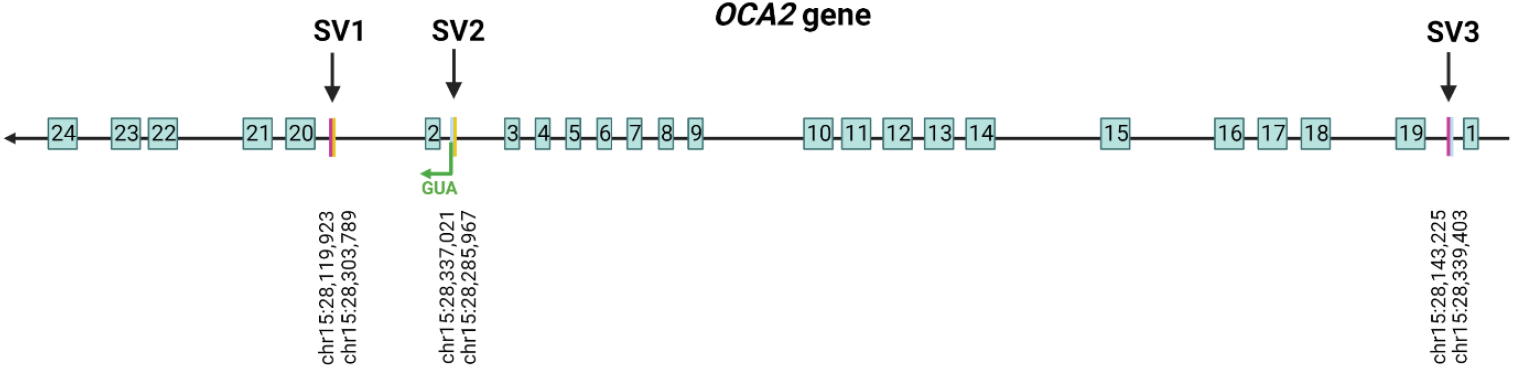
Reshuffled sequence of *OCA2* gene with primer positions indicated.

**Supplementary Figure 2:**
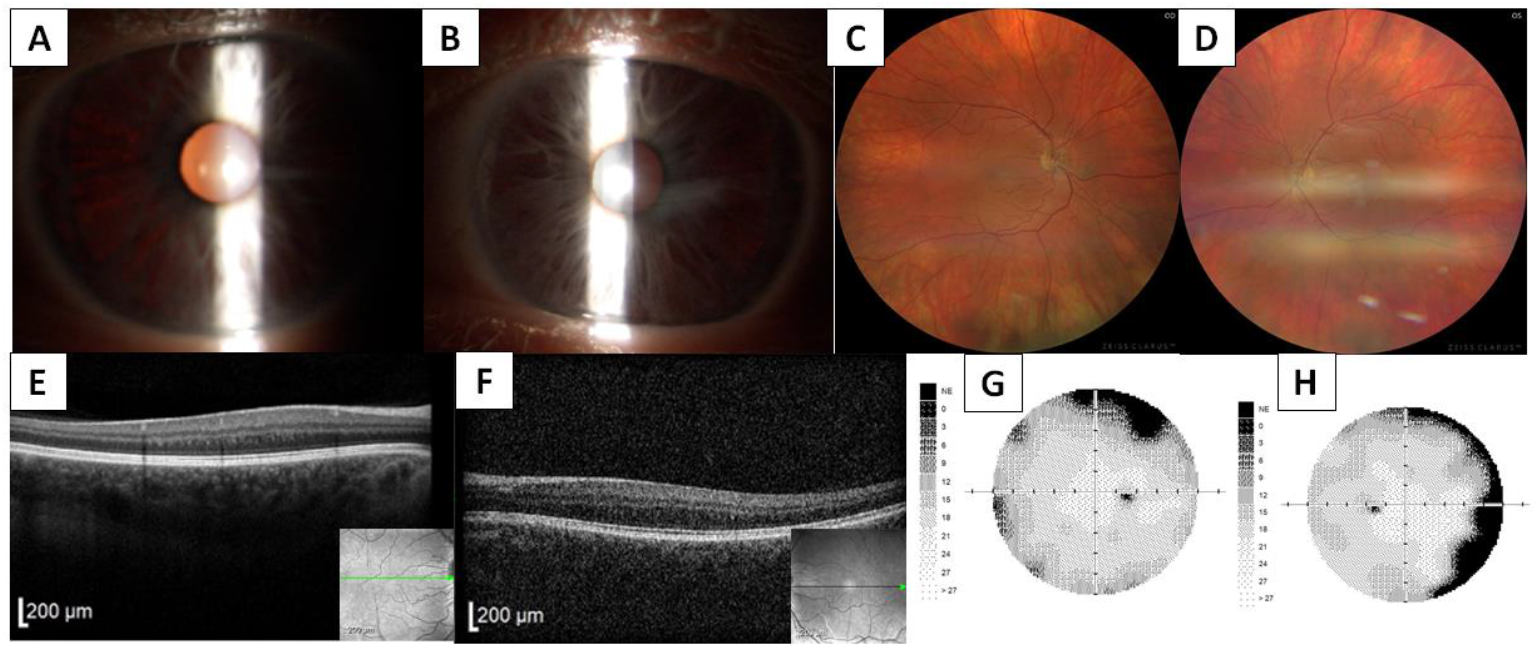
Clinical findings in individual II:3 with oculocutaneous form of albinism. Anterior segment photographs document iris translucency in the right (A) and in the left eye (B). Markedly hypopigmented fundus in the right (C) and in the left eye (D), absence of a foveal pit in SD-OCT scans of the right (E) and left eye (F) (horizontal scan line is provided in the inserts). The visual field in the right (G) and left (H) shows a mild non-specific decrease of sensitivity.

**Supplementary Figure 3:**
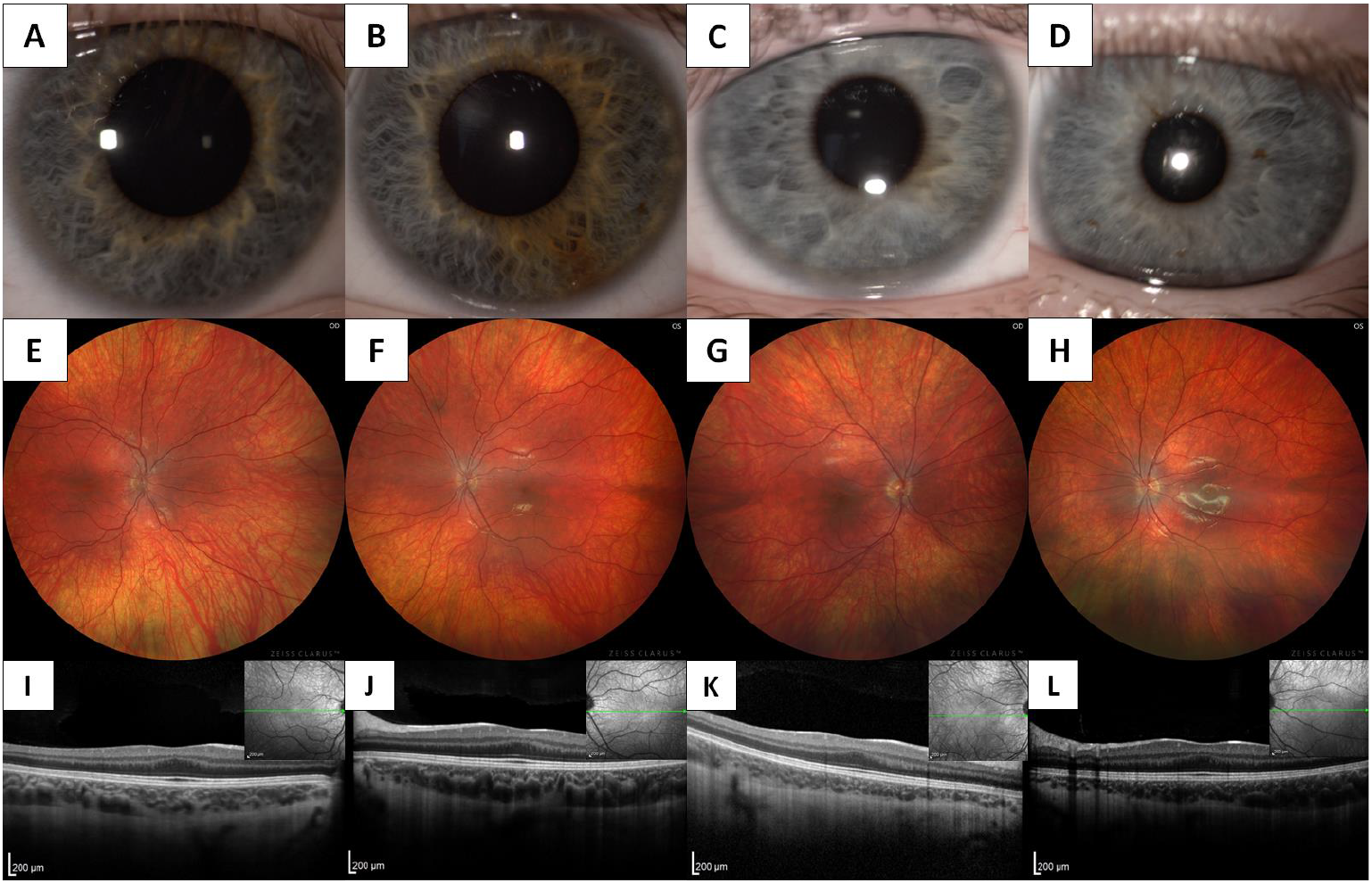
Clinical findings in two family members with an ocular form of albinism. Individuals III:2 and III:3, respectively, have normal anterior segment appearance of the right (A, C) and left (B, D) eyes, Wide-field fundus photography revealed discreet fundus hypopigmentation in the right (E, G) and left (F, H) eyes. Foveal hypoplasia (grade 1 - shallow foveal pit) was detected by SD-OCT in the right (I, K) and left (J, L) eyes (a horizontal scan line is provided in the inserts).

